# The anatomy of obsessive-compulsive disorder

**DOI:** 10.1101/2022.10.06.22280808

**Authors:** Iain E. Perkes, Mustafa S. Kassem, Philip L. Hazell, George Paxinos, Philip B. Mitchell, Valsamma Eapen, Bernard W. Balleine

## Abstract

OCD has been characterized by recent data as a disorder of cognition. Recent data also show pathology in prefrontal-subcortical networks. We leveraged cross-species prefrontal-subcortical cytoarchitectonic homologies in order to parse anatomical abnormalities in people with OCD into higher resolution areas and neuronal networks. We established that the anatomical abnormalities associated with OCD predominantly reside in a neuronal network associated with emotional processing. We further provide evidence that current tests do not accurately dissociate emotion from cognition and so relying on them risks mis-stating the role of prefrontal-subcortical networks. Taken together, these findings reveal the neglect of the role of emotion in the pathophysiology of OCD.

**Background:** Recent advances in the cytoarchitectonic parcellation of the human brain have significant implications for major psychiatric conditions such as obsessive-compulsive disorder (OCD). Brodmann’s areas have remained in use as the histological map of the human brain, framing its functional correlates in health and disease. However, cytological research has continued to refine these divisions in some cases substantially. For instance, the 16 areas in Brodmann’s prefrontal cortex have expanded to 63, delivering a four-fold increase in granular resolution. These contemporary cytoarchitectonic areas have been parcellated into distinct prefrontal-striatal networks responsible for (i) the control of emotions and visceral organs, (ii) mental representation and classification of external objects, and (iii) the control of visual attention. Interacting pathology across prefrontal-striatal circuits makes OCD a paradigmatic condition upon which to apply these advances. The enhanced granular and network resolution this provides could transform human brain imaging from the original divisions of 1909 to higher resolution delineations, for example, providing precise mediolateral partitioning of the orbitofrontal cortex, thereby distilling the substrates of obsessions and compulsions.

**Advances:** Here we provide a meta-review of existing reports of thousands of people with OCD to reveal impairments spanning sensory integration, affective arousal, cognitive control, and motor action selection. Behavioral data previously interpreted as implicating only cognitive abnormalities have failed to detect cognitive impairment in children and adolescents with OCD casting doubt on the sensitivity of conventional tests and the temporal relationship between apparent pathology in adults and OCD symptoms. Therefore, by relying on that behavioral evidence alone we risk mis-characterizing OCD solely as a disorder of cognition. Moreover, the presence of sensorimotor and neuroimaging abnormalities in young people with OCD indicate the chronological primacy of undifferentiated abnormalities in neuronal structure and function. Neuronal correlates of OCD symptoms were found to map evenly into emotional-visceral and object assessment networks; within the visual attention network only the premotor cortex had substantive abnormality. Tasks reported as measuring cognition also distributed equally across networks further calling into question the physiological fidelity of these tasks. In contrast, tasks reported as measuring emotion mapped faithfully onto the emotional-visceral network. Volumetric changes in people with OCD also implicated the emotional-visceral network, in which the number of abnormalities were double those in the object assessment network.

**Outlook:** Although conventional behavioral tasks characterize OCD as a cognitive disorder, associated anatomical abnormalities are, in fact, distributed across two distinct neuronal networks responsible for (i) the control of emotions and visceral organs and (ii) the representation of external objects. The predominance of abnormalities in an emotional-visceral neuronal network contrasts with the paucity of research on emotional processing in OCD relative to tasks reported to test cognition, showing an inflated attribution of cognitive relative to emotional dysfunction in the pathophysiology of OCD. The histologically derived orbital and medial prefrontal cortex subregions, shown here as selectively affected in people with OCD, provide higher resolution candidate treatment targets for neurostimulation and other therapeutics. Extending our current work to other conditions could identify transdiagnostic neural signatures of psychiatric symptoms.

**One-Sentence Summary:** Structural brain changes in people with OCD reside predominantly in a neuronal network responsible for emotional control.

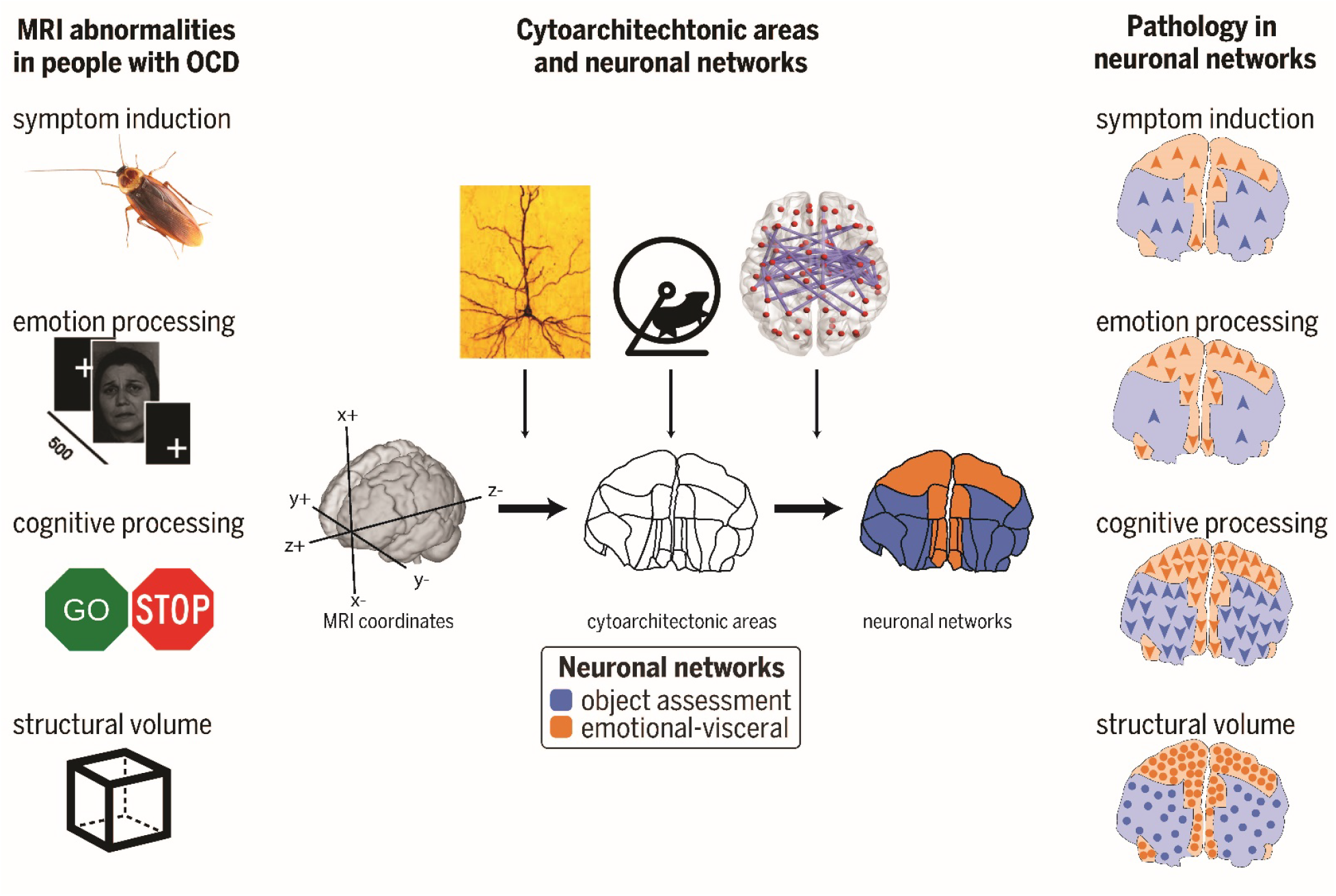

**OCD as a pathology of cytoarchitecture:** Neuronal networks derived from cross-species studies of cell structure, projections, and function transform the granular resolution of human brain imaging analysis to reveal the role of an emotional-visceral network in the pathophysiology of OCD.

## Main Text

Despite psychiatric disorders affecting over one billion people annually, research has failed to generate significant advances in treatment in recent decades (*1, 2*). There have been well publicized attempts to re-focus enquiry into the mechanisms of mental illness (*2*). However, ongoing structural issues impede the integration of basic and clinical neuroscience. Here we address one of these barriers through the lens of pathophysiology in OCD, capitalizing on its relatively clear characterization among psychiatric disorders and further warranted by the high disease burden (*3*).

In this context, recent technological developments provide promise; particularly those in neuroimaging, deep transcranial magnetic stimulation (TMS), and deep brain stimulation (DBS; Fig. 1A) (*4, 5*). Neuroimaging has outpaced other lines of investigation to reveal interacting pathology across prefrontal-striatal circuits in OCD (Fig. 1B). Nevertheless, neural circuit abnormalities are all too commonly interpreted using the large macroscopic areas, i.e., gyri and sulci or functional areas of older human atlases, i.e., Brodmann’s areas. Moreover, constructs such as the orbitofrontal cortex (OFC) combine areas that are heterogenous in function and cellular structure (*6*). As a consequence, there is an opportunity to enhance the resolution of these data by integrating recent anatomical advances to generate more accurate histological maps of neuroimaging from people with OCD (Fig. 1C). Macroscopic areas typically take their boundaries from cortical surface landmarks, however, some of these morphological areas, e.g., the OFC, are less clearly defined and have proven to be ineffective therapeutic targets for OCD. For example, TMS targeting the OFC has failed to ameliorate symptoms (*7*), whereas targeting the anterior cingulate cortex, prefrontal cortex, supplementary motor area—of which there is greater functional and structural resolution—have yielded positive results. Subcortical structures such as the nucleus accumbens and subthalamic nucleus, have shown effectiveness as DBS targets (Fig. 1A) (*8*).

**Fig. 1.**
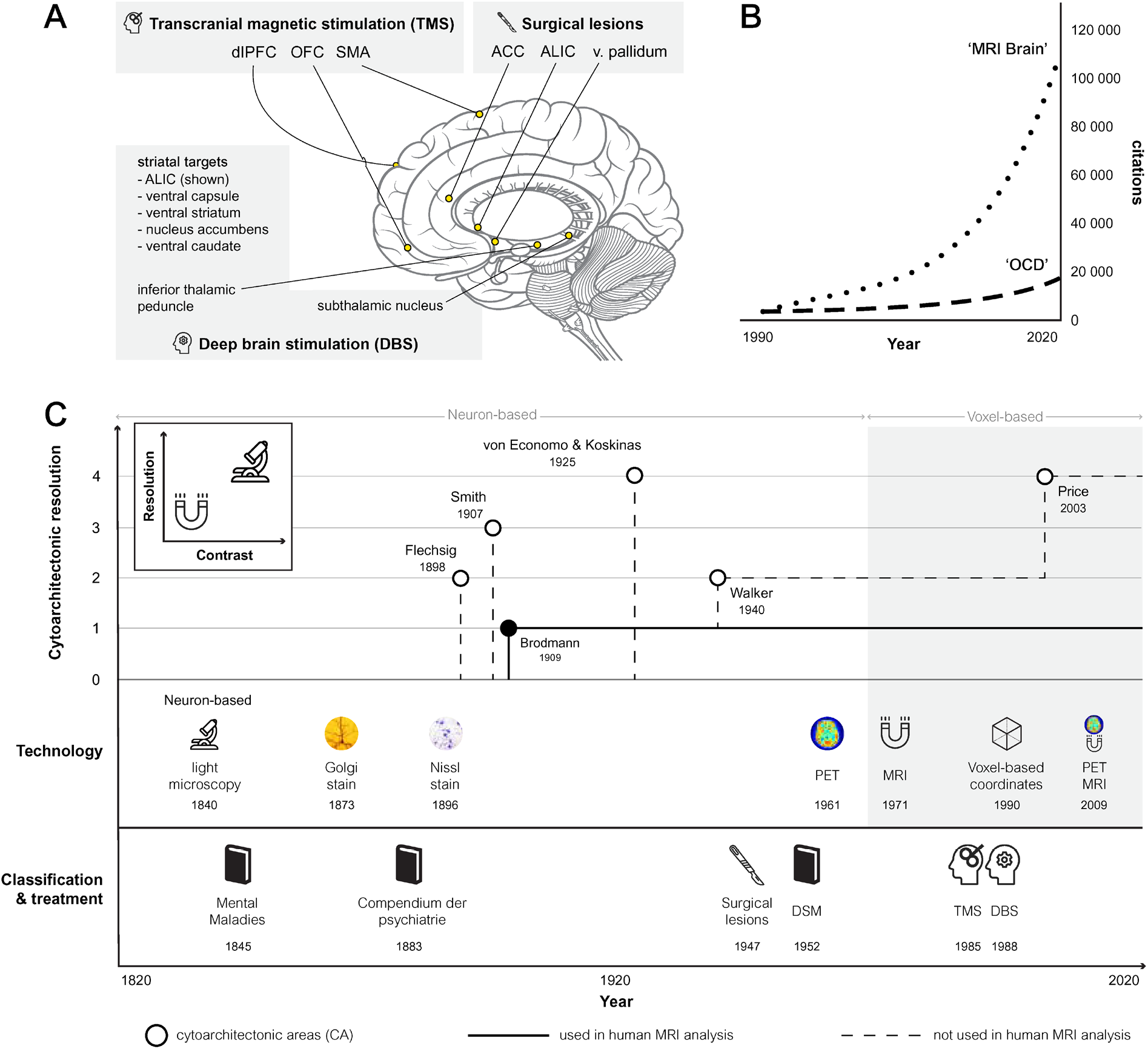
Localization in the cerebral cortex based on neurons and voxels. (**A**) shows therapeutic targets for TMS, historical surgical lesion studies (*9, 10*), and DBS. TMS is effective whether low (inhibits) or high (enhances cortical activity) frequency stimulation is applied. The supplementary motor area proved superior to the right dorsolateral prefrontal cortex (dlPFC), left dlPFC, or bilateral dlPFC (*4*); however OFC targets did not prove effective (*7*). For DBS, no difference in effectiveness was found between striatal and subthalamic targets, however DBS-responders had a greater age of onset (*8*). (B) shows the growth of in-title ‘OCD’ AND ‘MRI’ AND ‘Brain’ Pubmed-listed publications. *Panel C* shows (i) technologies used for brain parcellation ranging from the accessibility to light microscopy (*11*), Nissl staining (*12*), and Golgi staining (*13*) which enabled neuron-based delineations—Brodmann’s areas—later applied to the voxel-based technologies of MRI and positron emission tomography. (C) Classification in psychiatry has developed in parallel. Early diagnostic classifications were based on observation and description as opposed to the more technical approach that prized reproducibility in modern diagnostic manuals (*14*). In addition to the contrast provided by stains described above, modern microscopy provides less than 1-micron resolution (*15*) whereas MRI currently can reach 50-micron resolution post-mortem.

Part of the issue is precedence. Histological investigations of the structure and function of the brain were standardized as Brodmann’s areas in 1909. These have been retained in their unadulterated form in subsequent maps to frame functional correlates in health and disease. This is despite continuing research delivering a four-fold increase in resolution that incorporate connectivity and functional data across species to generate distinct prefrontal-striatal networks (Fig. 2) (*16-20*).

**Fig. 2.**
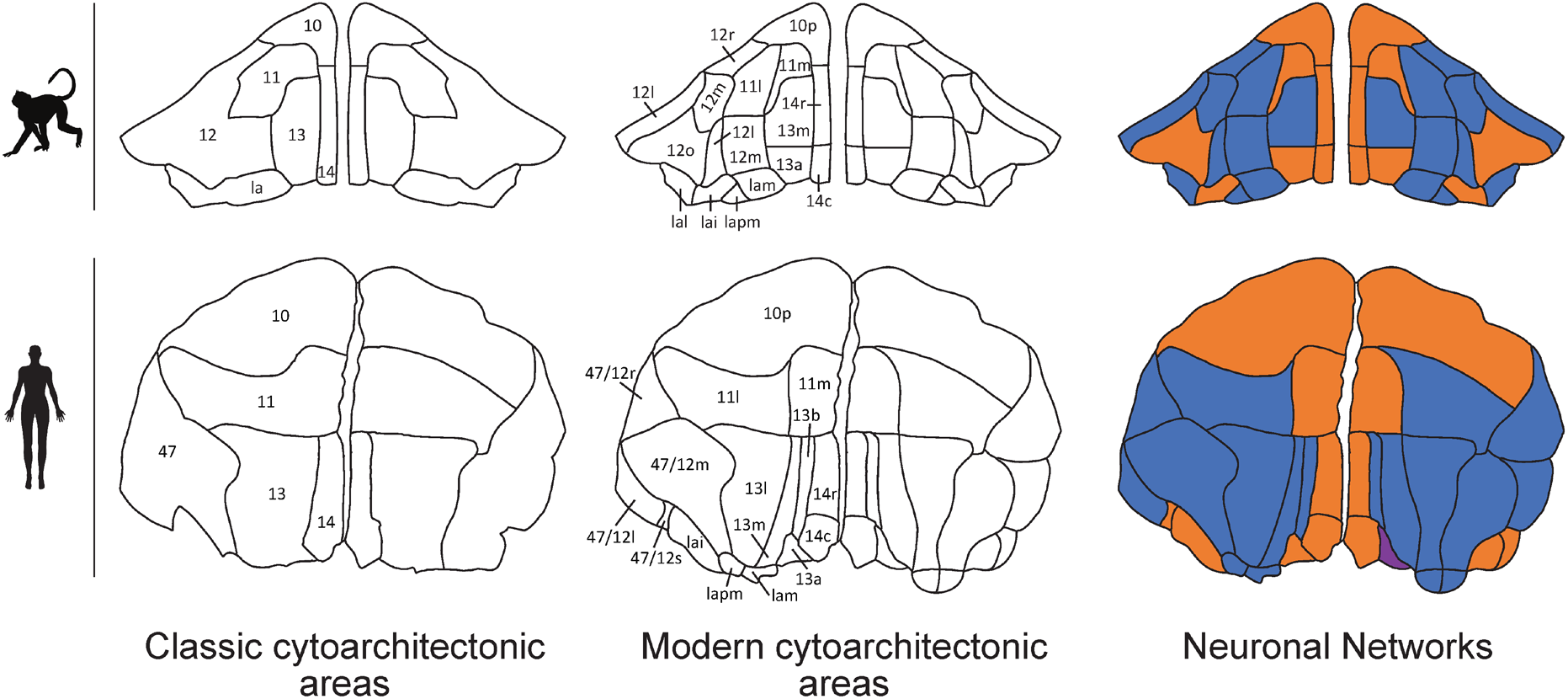
Cytoarchitectonic areas and neuronal networks in macaques and humans. Prefrontal parcellations and homologies between macaques and humans shown by transverse images of the OFC, progressing from classical, e.g., Brodmann’s Area 11 (BA11), to modern cytoarchitectonic areas, e.g., 11l (lateral), and then the neuronal networks responsible for object assessment (orange) and emotional-visceral control (blue). The object assessment network integrates multi-modal sensory input to assess identity, value, and expectancy of objects and guide decision-making. The emotional-visceral network processes emotion and controls viscera, i.e., autonomic and endocrine. It overlaps functionally and structurally with the ‘default mode network’ in that it is active during rest and introspection as occurs during ‘resting state’ functional MRI (fMRI) paradigms but is less active during externally directed tasks. Individual components of these networks are shown in Figure 5

We collected voxel-based MRI coordinates from meta-analyses of people with OCD and projected them onto recent brain maps derived from cross-species histological studies to provide a higher resolution picture of OCD. This resulted in a set of proposals about the functional changes in the architecture of the brain associated with OCD that provide a means of reconciling recent, sometimes quite disparate, claims regarding the neural bases of OCD.

### Behavioral effects of OCD

Recent attempts to provide objective functional descriptions of psychiatric disorders have reduced confidence in more subjective diagnostic approaches (*2*). The restriction to symptom-based diagnosis differentiates mental illness from other medical conditions where physical examination, radiological, and biochemical findings are incorporated. This differentiation represents the limits to our knowledge of the mechanisms of mental illness but does not negate diagnostic reliability or validity (Fig. 1C). The symptoms of OCD, for example, have considerable construct validity. A recent meta-analysis of symptoms from over 5000 people with OCD confirmed the identity of symmetry, taboo, cleaning, and hoarding as the archetypal symptom domains and the stability of these domains neurodevelopmentally and across cultures (*21*). Furthermore, there were behavioral abnormalities across sensory, motor, affective, and cognitive functional domains.

With regard to sensory processing, it is well understood that, in OCD, sensory stimuli may precipitate obsessions and compulsions and, in tests of sensory integration, when asked to identify standardized external stimuli (e.g., a figure-of-8 traced onto the palm of the hand), meta-analyses showed impaired ‘sensory integration’ (*22*) (Fig. 3). Motor difficulties in OCD, also confirmed by meta-analyses, include motor coordination, speed and planning (*22-25*). Additionally, dysregulation in innate and conditioned sensorimotor reflexes, e.g., ‘primitive’ reflexes, such as sucking and palmomental arcs (*22*), often resist descending cortical control during neurodevelopment to persist in adults with OCD (*22*) and so provide one broad marker of neuropathology. Furthermore, voluntary control of innate visual gaze reflexes, assessed by asking people to suppress the reflex to look towards novel stimuli in the anti-saccade test (*26*), is diminished in people with OCD suggesting a deficit in the control of saccade-generating neurons of the superior colliculus (*26*). Nevertheless, as with many findings in OCD, it is not known whether these impairments in the automatic and voluntary control of innate reflexes precede or follow the onset of OCD and, therefore, whether they are a cause or consequence of the illness. Such deficits in volitional control extend to cognitive domains, with repetitive behaviors related to impaired suppression of prepulse motor activity and disinhibition demonstrated by ‘go/no-go’ and related tasks, e.g., ‘updating’ or ‘set-shifting’, providing evidence of mental rigidity (*23-25*). However, as the complexity of behavioral tasks increases, more mental functions, e.g., attention, memory, secondary (conditioned) motivation, are enlisted and the functional and anatomical interpretation accordingly becomes more nuanced. Indeed, despite early attention from psychodynamic theories generally, the empirical evaluation of emotional changes in OCD is under-explored. We identified only one meta-analysis focused on emotion in people with OCD relative to healthy controls that revealed ‘intolerance uncertainty’, although this marker does not isolate emotion and it did not differentiate people with OCD from those with generalized anxiety disorder or major depressive disorder (*27*). However, insomnia is a broad marker of over-arousal and has been confirmed in people with OCD relative to healthy controls (*28, 29*).

**Fig. 3.**
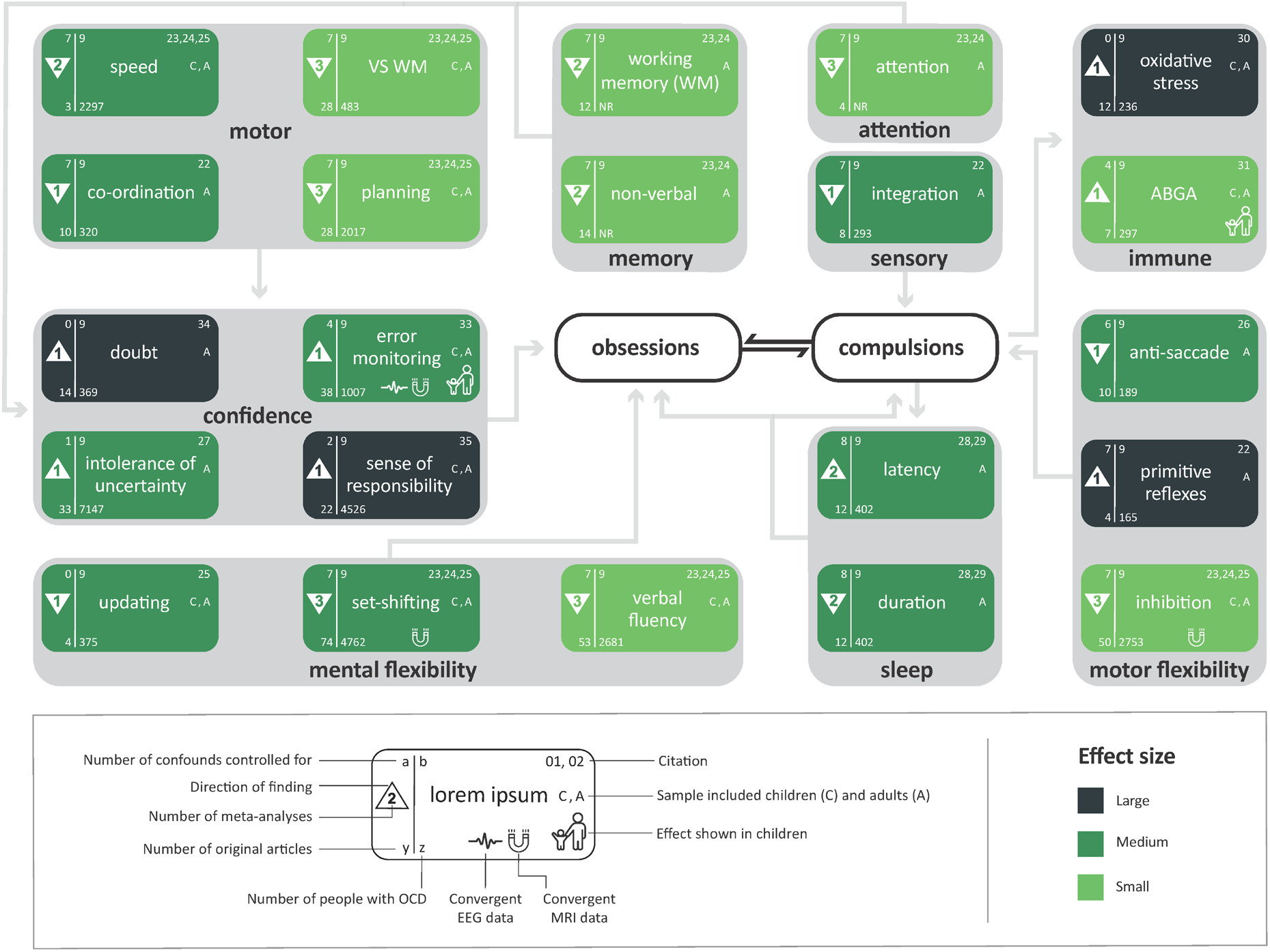
Behavioral changes and putative interactions in OCD. Pooled results of meta-analyses of behavioral abnormalities in people with OCD relative to healthy controls are presented by effect size, sample type, power, and control of confounds (age, medication, comorbidities, gender, education level, age of onset, duration of illness, OCD symptoms, and depressive symptoms). Results (except sleep) survived control for age, medication, and comorbidities. Grey arrows represent speculative interactions. Performance intelligence was incorporated into motor speed. Conflicting results are not displayed, e.g., cytokines. Visuospatial working memory (VS WM) also denotes non-verbal working memory impairment. Working memory denotes visual and verbal working memory. Error monitoring represents error-related negativity. ABGA=anti-basal ganglia antibodies; EEG=electroencephalogram.

Another area that appears under-explored is the clinical presentation of OCD in children and adolescents. This is critical because OCD has its onset during this period yet, surprisingly, meta-analysis does not show cognitive impairment in children with OCD (*32*). Indeed, the lone behavioral abnormality reported in children with OCD that survives meta-analysis is *error related negativity*, a proxy for action monitoring, measured as a change in EEG voltage after an erroneous response to a stimulus (*33*). Related constructs such as ‘increased sense of responsibility’ and ‘doubt’, provide further evidence of the increased salience of getting things ‘right’, or ‘just so’, in people with OCD (*34, 35*). These concepts relate to the suggestion that doubting/checking phenomena are related to the (false) belief that a given action will prevent a (generally unrelated) outcome; something that has led to the investigation of inflated perceived agency via thought-action fusion (*36*).

This pattern of behavioral deficits introduces a focus on distinct mechanisms of obsessive and compulsive behaviors. The occurrence of some level of obsessions or compulsions in around 80% of people without OCD, together with the stability of obsessive-compulsive symptoms across cultures—in contrast to other psychiatric symptoms, e.g. delusions; (*37*)—forces the consideration of OCD pathophysiology as involving excess generation (hypergenesis) and insufficient termination (hypodismissal). That people with OCD had greater symptom inducibility compared to healthy controls supports the hypergenesis mechanism, which is generally argued to reflect excessive affective arousal, motivating ‘harm avoidance’ and ‘incompleteness’ (*38*). However, such interpretations are limited by the paucity of research into emotion processing in OCD. In contrast, hypodismissal may be due to impaired inhibitory mechanisms, e.g., in impaired extinction learning and cognitive control. Supporting this hypothesis, a cluster of impairments under the rubric of cognitive flexibility—impaired set-shifting (*23-25*), verbal fluency (*23-25*), and updating (*25*)—may help explain why people with OCD are ‘stuck’ in symptom loops. Similarly, motor rigidity—disinhibition (*23-25*), primitive reflexes (*22*), and reflexive eye movements (*26*)—may relate to compulsion hypodismissal. Other motor control impairments—incoordination, slowness, disinhibition, visuospatial working memory, and performance IQ—may also relate to compulsion hypodismissal in the broad sense that they all control motor performance. This motor theme is supported by the fact that the supplementary motor area, responsible for motor planning (*39*), is the most effective TMS treatment target for OCD (*4*). The relatively selective impairment in non-verbal, as opposed to verbal, working memory may also reflect impairments in performance, rather than in ‘thinking’ (*23*). Likewise, reduced sleep diffusely compromises cognitive and behavioral control and so limits dismissal of both obsessions and compulsions (*5*).

In summary of behavioral data, we see a 10:1 ratio of ‘cognitive’ against ‘emotional’ meta-analyses, a ratio that balloons to 21:1 when considering the individual tasks, e.g., set-shifting, within those studies. These data also show signals of rigidity and doubt in addition to several sensorimotor abnormalities. Indeed, even for those tasks that have been examined using fMRI, e.g., inhibition, motor planning, and set-shifting, the use of analytic methods of either Brodmann’s original areas or heterogenous macroscopic regions limits the resolution of the map to which these functions are associated.

### Beyond Brodmann: The neural effects of OCD

Brodmann’s 1909 mapping of the human cortex into 52 areas (*40*) drove considerable subsequent research in brain architecture (Fig. 1C) (*41*). More recently, Price has further delineated Brodmann’s and Walker’s areas, within the PFC, to reveal an additional 22 areas using histochemical staining to characterize the cytology, myelination, metabolism, neurotransmission, and neuronal projections of the macaque brain (*19*) (Fig. 2). These 22 areas were then translated from the macaque to human before they, alongside other cytoarchitectonic areas, were aligned into three networks based on their functional and structural fidelity (*16-18*).

To enable a more contemporary investigation of OCD pathophysiology, we clarified and standardized the high-resolution histological data that Price and others illustrated in their studies of human and animal brains (Methods). In addition to this general cortical remapping, the recent delineation of the OFC into 19 areas based again on cytoarchitectonic data, is of relevance given recent evidence of orbitofrontal changes in people with OCD (*42, 43*). Our work aligned these 19 orbitofrontal areas into three regions—the medial edge, central mass, and caudolateral patch— based on cross-species histological and functional data. These three regions then independently aligned with other cortical and striatal regions to constitute neuronal networks associated with functions that are characterized as ‘object assessment’, ‘emotional-visceral control, and ‘visual control’. For example, the orbitofrontal medial edge aligned with the medial caudate and ventrolateral PFC as part of the emotional-visceral neuronal network. BA47 shows the benefit of this re-organization in that it subdivides, based on its cellular content, into 47/12r, 47/12m, 47/12l and 47/12s. Area 47/12s, for instance, has more connections in common with the intermediate agranular insula (iai) than with areas 47/12r, 47/12m, or 47/12l. This unique connection between 47/12s and iai aligns with the emotional-visceral network, whereas the remainder of BA47 aligns with the object assessment network.

A defining characteristic of the object assessment network is its receipt of inputs from cortical areas associated with sensory systems, e.g., vision, audition, smell, taste, and visceral afferents including somatic sensation (*20, 44-47*). The broad function of this network is, therefore, considered to be the integration of these sensory inputs to determine the identity of external objects. The object assessment network consists of the OFC central mass, ventrolateral PFC, lateral caudate, and medial putamen (*16, 18-20*). As distinct from the emotional-visceral network there are few known outputs from the object assessment network to the midbrain.

In contrast, the emotional-visceral network has few sensory inputs and is instead characterized by outputs to visceral control structures such as the midbrain, including the periaqueductal gray and both medial and lateral hypothalamic nuclei, indicating coordination of autonomic and endocrine function (*17, 18*). The emotional-visceral network includes the OFC medial edge, OFC caudolateral patch, dorsolateral PFC, hypothalamus, majority of the cingulate cortex and hippocampus, the rostromedial caudate, and ventral putamen.

The visual attention network consists of three cytoarchitectonic areas in the caudoventral, caudolateral, and premotor PFC regions (*18*). Its primary role is thought to be the control of saccadic eye movements (*17, 48*); it may also be part of the dorsal attention system (*18*).

### A metanalytical analysis of neuronal networks in OCD

To validate and clarify this approach we used these cytoarchitectonic areas and neuronal networks to analyze data arising from meta-analyses derived from people with OCD that included the description and interpretation of behavioral abnormalities (see Methods).

Symptom activation procedures, such as watching a video of cockroaches, were found to have provoked neuronal hyperactivity in a circuit involving the OFC and putamen (Fig.4; Fig 5).

**Fig. 4.**
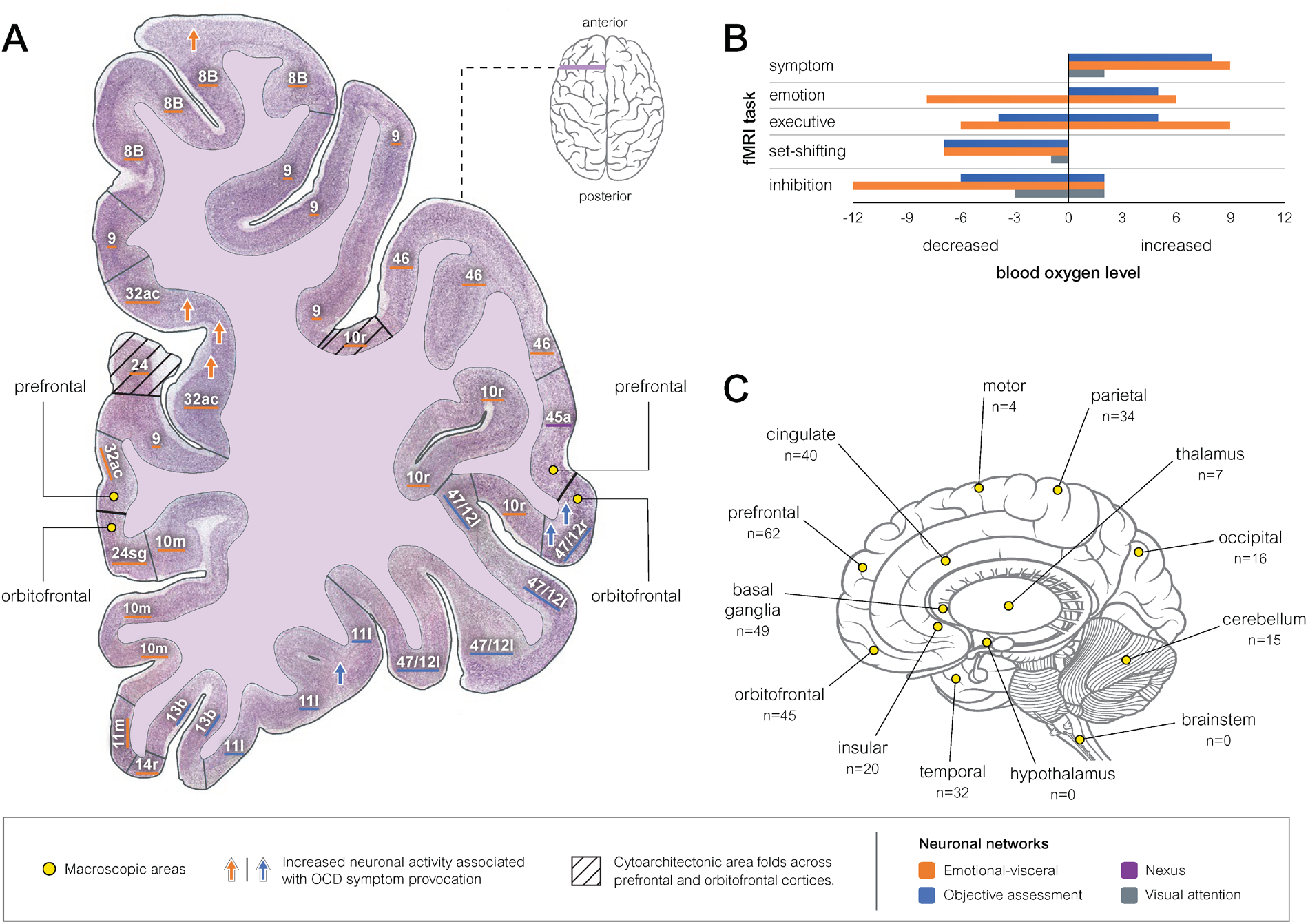
Classical and contemporary parcellations of the prefrontal cortex. (**A**) demonstrates modern cytoarchitectonic and neuronal networking analysis by projecting 7 of the 19 symptom-provocation foci onto a Nissl-stained coronal histological plate to reveal that even within the ‘OFC’ these results are distributed across networks. (B) shows blood oxygen level dependent (BOLD) abnormalities in people with OCD compared to healthy controls during task-based fMRI; these 150 data points are drawn from 10 meta-analyses of people with OCD compared to healthy controls. (C) shows all OCD abnormalities from our review of meta-analyses, grouped by conventional anatomical verbal descriptors. Any single conventional region, e.g., PFC, accounted for only a minority of (n=62/332) abnormalities.

**Fig. 5.**
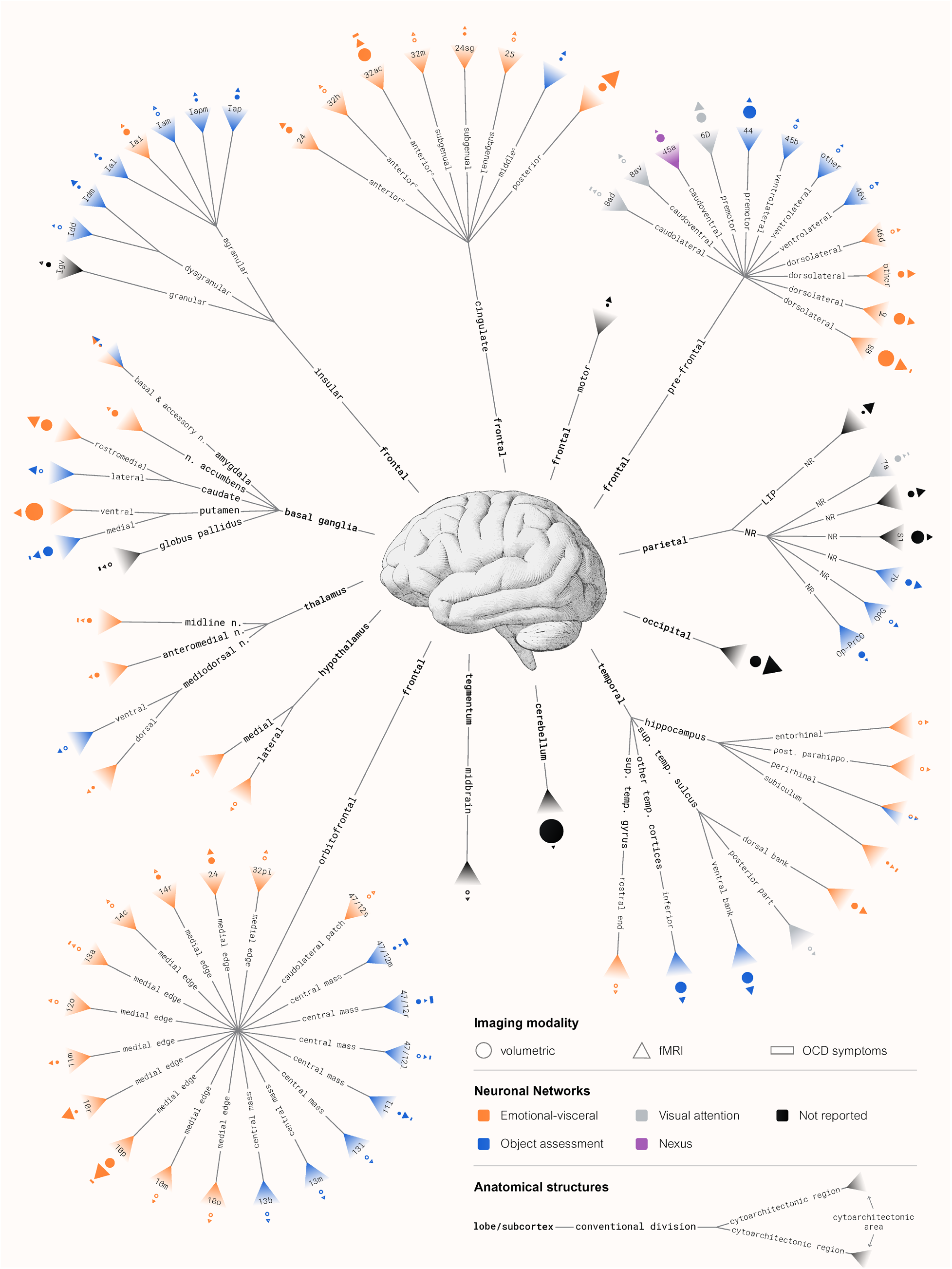
Cytoarchitectonic areas, neuronal networks, and OCD. Cytoarchitectonic area and neuronal network differences in people with OCD during volumetric, fMRI symptom induction and other fMRI imaging compared to people without OCD represented by shape- and color-coded symbols adjacent to anatomical structures and sized proportionate to the number of abnormalities. The colors ‘black’ and ‘purple’ are non-networked and multiple networked areas respectively. Empty symbols denote nil abnormalities. Total volumetric abnormalities (n=170) were divided into the emotional-visceral (↑36: ↓36), object assessment (↑20: ↓16), and visual attention (↑5: ↓2) networks. General (non-symptom) fMRI abnormalities (n=140) were divided into the emotional-visceral (↑22: ↓37), object assessment (↑10: ↓20), and visual attention (↑4: ↓4) networks. Full anatomical resolution was restricted to networked cytoarchitectonic areas, consequently the depicted non-networked areas, e.g., cerebellum, are artefactually inflated in this graphic. Within a given cytoarchitectonic areas nil abnormalities in one imaging modality, e.g., volumetric, predicted nil abnormalities in the other imaging modalities, i.e., symptom induction and general fMRI; providing convergent evidence for this new method. For instance, 47/12s had nil abnormalities across all modalities whereas 47/12m had abnormalities all MRI modalities. The 19 symptom induction abnormalities were within the OFC central mass (n=6), OFC medial edge (n=3), anterior cingulate cortex (n=3), dorsolateral (n=1) and caudolateral PFC (n=1), parietal lobe (n=1), medial putamen (n=2), midline nucleus of the thalamus (n=1), and subiculum (n=1).

However, the PFC predominated with respect to volumetric and general fMRI abnormalities in people with OCD. Neuronal network analysis showed a general 2:1 distribution of abnormalities between the emotional-visceral and object assessment networks, consistent across cytoarchitectonic regions within conventional areas; e.g., the OFC medial edge relative to the central mass (Fig. 5).

OCD symptom induction abnormalities were localized to an orbitostriatal circuit consisting of the OFC medial edge, OFC central mass, anterior cingulate cortex, and ventral putamen. Abnormalities in neuronal activity (BOLD) were uniformly increased during symptom provocation tasks. This excess neuronal activity is consistent with ‘excess’ obsessions and compulsions, provocation procedures also induce some degree of obsessions and compulsions in people without OCD. Similarly, decreased BOLD during set-shifting and inhibitory tasks was associated with reduced behavioral performance in people with OCD (Fig. 4). BOLD can represent compensatory mechanisms (*49*); however, congruence in the direction of the behavioral and BOLD results supports the suggestion that these results are mediating, rather than compensating for, symptoms and behavioral function respectively.

OCD symptom activation occurred in similar proportions across the object assessment and emotional-visceral networks, indicating that obsessions and compulsions are associated with both cognitive and emotional difficulties. The remaining two of the 19 abnormalities were in the visual attention network; therefore, all OCD symptom foci were in the three defined neuronal networks, supporting the use of these networks to investigate OCD pathophysiology.

OFC central mass-based symptom abnormalities were further concentrated in BA47, an area linked to semantic language disambiguation and processing of emotion on human faces (*50*). Converging with this BA47 anatomical abnormality, intolerance of semantic uncertainty has been shown in a meta-analysis of 7147 adults with OCD (*27*). However, as with other human imaging studies, these functions were not divided by BA47 subdivisions.

Conventional area abnormalities for volumetric and general fMRI modalities in the PFC were similar in number to those in the OFC (Fig. 4). However, symptom induction abnormalities in the OFC outnumbered those in the PFC 9 to 2; suggesting that the PFC mediates the sequelae of OCD, whereas the OFC and anterior cingulate cortex mediate the symptoms. Nevertheless, supporting the suggestion that volumetric and general fMRI abnormalities reflect the neuronal sequelae of OCD, 18 of the 19 symptom induction studies were associated with cytoarchitectonic areas that also had abnormalities in both volumetric and fMRI modalities.

We used an analysis seeded from the orbital and medial prefrontal cortex (OMPFC), this was supported by its predominant capture of OCD abnormalities. Across all fMRI abnormalities, 86% (n=144/168) were accounted for by the three neuronal networks. Across all volumetric abnormalities, 82% (n=140/170) were accounted for by the three networks; of the remainder, changes in the cerebellum accounted for nearly half (n=14/30). This unanticipated cerebellar pathology in OCD converged with abnormalities in other areas relating to coordination and control of motor output—the caudate, putamen, and premotor cortex (particularly areas 44, n=9, and 6D, n=10; Fig. 5).

The putamen contained more abnormalities than the caudate across all imaging modalities, consistent with reports of cerebrovascular events in the putamen and globus pallidus associated with the generation of repetitive movements (*51*) and with OCD in humans (*52*). Nevertheless, this relative difference between the caudate and putamen was surprising given studies showing modulation of caudate volume in response to pharmacological and behavioral therapies (*3*). Whereas OCD symptoms were associated with a 1:1 ratio between the emotional-visceral and object assessment networks, a 2:1 pattern emerged in volumetric and general fMRI in people with OCD relative to those without the condition. This predominance in the emotional-visceral network across the whole brain was consistent within conventional regions, e.g., the OFC and caudate (Fig. 4). For instance, OFC medial edge (emotional-visceral network) predominated relative to the OFC central mass (object assessment) in terms of volumetric and general fMRI abnormalities.

Evaluating the brain structures whose functionality is spared by OCD is equally important in understanding the disease. The ventrolateral PFC, responsible for decision-making under ambiguity (*53*), showed no abnormalities in people with OCD suggesting that it is not decision-making *per se* that is disrupted in people with OCD but rather its emotional sequelae. The hypothalamus, associated with visceral control (*54*), also showed no abnormalities—a salient finding given that the emotional-visceral network was otherwise strongly implicated, perhaps indicating that it is emotional processing, rather than visceral control, that is disrupted in people with OCD. Despite suggestions that the lateral OFC is hyperactive in OCD (*43*), the OFC central mass (which includes the lateral OFC) showed relatively few volumetric or general fMRI abnormalities (Fig 4). The lateral caudate showed no volumetric, and few task-based, abnormalities. The hippocampus—associated with spatial memory, navigation, and emotional behavior—showed aberrance only in the subiculum which is the main output region. This is also surprising when set against evidence of memory and general visuospatial deficits in people with OCD (*23-25*).

### The bias in cognitive vs. emotional abnormalities in OCD

Meta-analysis of classical behavioral test results in people with OCD reveal a 21:1 preponderance of abnormalities characterized as ‘cognitive’ relative to ‘emotional’, giving rise to the concept of OCD as a disorder of cognition (*55*). We therefore expected that the object assessment network would be selectively affected. In contrast, we found a two-fold predominance of abnormalities in the emotional-visceral network. This striking difference between behavioral and anatomical results may have been driven by the trend of the field toward ‘cognitive’ testing in addition to the limited precision of the tests themselves. Indeed, here we showed that, despite being framed as ‘cognitive’, task-based fMRI abnormalities in people with OCD parse most clearly into emotional-visceral and object assessment neuronal networks in equal measure (Graphical Abstract). For example, disinhibition, shown in three well-controlled meta-analyses of 2753 people with OCD, is commonly argued to be mediated by BA44, and yet, as reported within the meta-analyses, the 25 abnormalities in inhibition task-related fMRI in people with OCD were diffuse—there were no more than two foci within a single cytoarchitectonic area (Fig. 4). Within the visual attention network only the premotor (area 6D) had substantive abnormality. ‘Emotional’ task-based fMRI results, on the other hand, mapped onto the emotional-visceral network (Graphical Abstract) (*56, 57*).

In addition to limited precision, classical behavioral tasks may also be insensitive. A paradox casting doubt over the causative role of cognitive impairment is its apparent absence in children with OCD (*32*) despite meta-analyses replicating cognitive impairment in adults with the condition (*23-25*). Tasks that are less reliant on verbal instruction show promise here. For example, error-monitoring is the sole abnormality that survives meta-analysis in children with OCD. Other tasks with minimal verbal instruction, e.g., anti-saccade, sensory integration, and primitive reflex assessments, showed medium-to-large effect sizes in adults with OCD by well-controlled meta-analyses yet are underexplored in children with OCD.

### The constantly evolving parcellation of the brain

Recent comparative anatomy is incorporated and available in brain atlases, enabling the immediate implementation of reporting human imaging results against modern cytoarchitectonic areas (*42*), yet all of the eighteen MRI meta-analyses included here reported results using Brodmann’s original areas. We selected an OMPFC-focused networking approach because of the evidence that the OFC is selectively affected in OCD. We were, nevertheless, agnostic in our systematic approach to reviewing all the evidence for MRI abnormalities associated with OCD. Our non-networked anatomical analyses were also theory-neutral and unaffected by the OMPFC focus—these analyses showed that, while the OFC is indeed paramount in mediation of OCD symptoms, the PFC is equally affected in terms of structural changes. Moreover, there are other substantive abnormalities in the anterior cingulate cortex and putamen in addition to unanticipated findings in structures such as the cerebellum, occipital, and parietal cortices (Fig. 4). The cerebellar, basal ganglia, and premotor findings highlight a candidate role of motor control and execution systems in the pathophysiology of OCD.

Signs of behavioral inflexibility in classical tasks, e.g., set-shifting and inhibition, can be investigated more broadly to isolate the emotional drivers, cognitive controls, and neuronal substrates across species (*58*). Indeed, emerging evidence shows disruption to the balance between reflex and volition in people with OCD (*43*). Moreover, the measurability of compulsions makes OCD among the most penetrable psychiatric disorders with respect to cross-species behavioral neuroscience. For instance, compulsion-like grooming has been induced in rodents by optogenetic stimulation of corticostriatal circuitry and then rescued using medications known to be effective in treating OCD in humans (*59*). Meta-analysis of 4900 people showed that symptoms are inducible by standardized stimuli in healthy, subclinical, and clinical populations, albeit with different effect sizes between these groups (*60*). The symptom induction approach could readily be paired with cross species behavioral tasks and cytoarchitectonics in human studies.

OCD symptoms are mediated by hyperactivity across neuronal networks in a circuit consisting of the medial OFC, centrolateral OFC, anterior cingulate cortex, and medial putamen. Volumetric and resting state MR data selectively implicate the emotional visceral network (*61-70*). These methods show transdiagnostic promise—a meta-analysis comparing people with OCD to those with autism spectrum disorder, revealed that abnormalities concentrated in the emotional-visceral network (*65*), supporting the salience of the emotional-visceral network in, and providing a candidate biomarker for, OCD among neurodevelopmental disorders.

The hope is that adopting a more contemporary histology-based approach will finally shift human brain imaging analysis beyond Brodmann. Longitudinal application of cross-species tasks, combined with multi-modal imaging, across neurodevelopment could unlock the temporal sequence of pathology and symptoms in OCD. While such data are gathered, an extension of the current work to parse pathology associated with other psychiatric disorders will provide a transdiagnostic map of psychopathology in the brain. These higher resolution anatomical parcellations and specific behavioral analyses could transform precision medicine in psychiatry by refining neurostimulation and behavioral therapy targets in people with OCD.

## Methods

### Literature review

Inclusion criteria: (1) English language, (2) peer-reviewed original research or reviews that (3) informed the pathophysiology of OCD (pharmacotherapy trials excluded) (4) including human participants (5) with age reported and (6) diagnosed with OCD. Google Scholar, ScienceDirect, Web of Knowledge, Scopus, and PubMed searched on February 3, 2019 with no limits. Search terms: “obsessive-compulsive disorder” OR “OCD” AND “meta-analysis” OR “systematic review” in title. We manually searched reference lists and included articles known to us. We applied (7) a subtopic-specific evidence level criterion (systematic review [n=2] or meta-analysis [M-A; n=44]) to arrive at 46 publications, albeit excluding some interesting data (*71*). We discarded duplicates then classified findings by pre-defined subtopics to yield 46 (18 MRI, 10 cognitive, 5 neurostimulation or surgery, and 13 other) publications. Twenty-two MRI meta-analyses met inclusion criteria; four were excluded (*72-75*) (did not report anatomical coordinates), 18 underwent final analysis (*56, 57, 61-70, 76-81*). Peak voxels of 356 anatomical abnormalities (volumetric n=170; fMRI n=159; diffusion n=24) in people with OCD, compared with healthy or clinical control groups, were extracted from the 18 included meta-analyses (Fig. 5). Unless stipulated, clinical and demographic variables were controlled for by the source meta-analysis and did not moderate main effects. ‘Children’ refers to participants <18 years and ‘adult’ refers to those ≥18 years. We use the term ‘people’, or do not otherwise specify, when meta-analyses report a combined group of children and adults.

### Anatomical analysis

Diffusion foci (n=24) were excluded, this left 332 foci for analysis. Talairach coordinates transformed to Montreal Neurological Institute (MNI) coordinates (*82*). Each foci was dual visualized at 1.5T (1mm) (*82*) (https://bioimagesuiteweb.github.io/webapp/) and 7T (0.25mm) brain (*83*), using the MRIcron software (www.nitrc.org). The two resulting crosshairs were then both positioned in a modern atlas (*42*) for each foci to identify both (i) a physical structural feature (e.g., inferior frontal gyrus orbital part) and (ii) cytoarchitectonic area (CA; e.g., 11a). If the 1.5T and 7T crosshairs did not agree on a structure or BA, then the 7T crosshair was used. If a crosshair posited a structural border, then we referred to verbal anatomical descriptions in the source meta-analysis. This initial area-based allocation was first done at the highest available resolution; area-based allocation was then, where necessary collapsed to the Price level so that these CA could be allocated to a network. These morphology- and histology-based data were then compared to the subdivisions of the macaque monkey and human (*16, 18-20*). Each anatomical foci was assigned a subdivision e.g., BA11/subdivision 11a. For CA that do not translate as exact homologies, i.e., CA divided by the principal sulcus in the macaque, were assigned best-of-fit to the human. For example, subdividing BA8, there is a dorsal-most 8B, a dorsal to the principal sulcus 8ad and a ventral to the principal sulcus 8av. In this circumstance, as the human does not have a homology of the principal sulcus, BA8 was divided into three parts based on a dorsal most (8B), a ventral most (8av), and a middle portion that remained by subtraction of the other two (8ad). By dividing BA8 we developed a means that could best, without a homology, divide the corresponding BA. We analyzed neuronal network distribution patterns for imaging modalities with five or more abnormalities.

## Data Availability

All data are available online in the

## Acknowledgments

We thank Elijah Shervey for proof-reading and formatting, Faysal Ayub for discussion of the content, and Edmond Yang for assistance with design and preparation of graphics.

## Author contributions

Conceptualization: IEP, MK, PLH, GP, PBM, VE, BWB

Methodology: IEP, MK

Investigation: IEP, MK Visualization: IEP, MK

Funding acquisition: IEP, PBM, BWB, GP

Writing – original draft: IEP, MK

Writing – review & editing: IEP, MK, PLH, GP, PBM, VE, BWB

## Data and materials availability

On request.

